# Percent Thrombus Outperforms Size in Predicting Popliteal Artery Aneurysm Related Thromboembolic Events

**DOI:** 10.1101/2023.10.09.23296778

**Authors:** Tiffany R. Bellomo, Guillaume Goudot, Srihari K. Lella, Brandon Gaston, Natalie Sumetsky, Shiv Patel, Ashley Brunson, Jenna Beardsley, Nikolaos Zacharias, Anahita Dua

**Affiliations:** Division of Vascular and Endovascular Surgery, Massachusetts General Hospital, Boston, MA USA; Division of Cardiology, Noninvasive Cardiac Laboratory, Massachusetts General Hospital, Boston, MA USA

**Author notes:** Corresponding Author: Tiffany R Bellomo MD, Division of Vascular and Endovascular Surgery Massachusetts General Hospital 15 Parkman St, WACC-440, Boston, MA 02114. Authors contributed equally to the manuscript (Tiffany R. Bellomo, Guillaume Goudot).

**Keywords:** popliteal artery, aneurysm, ultrasound, non-invasive vascular lab

## Abstract

**Introduction:** Popliteal artery aneurysms (PAAs) are associated with high morbidity and mortality and current Society for Vascular Surgery (SVS) recommend operative repair for PAAs with a diameter greater than 20 mm, which is based on limited evidence. To help risk stratify patients for surgery, our aim was to identify anatomic characteristics of PAA associated with limb threatening and thromboembolic events (TEs).

**Methods:** A retrospective multi-institutional cohort was queried for all patients with a PAA from 2008 to 2022. Symptom status at the time of presentation was divided into three categories: symptomatic PAA with documented claudication or chronic limb ischemia (CLI), limb threatening PAA with a TE, acute limb ischemia, or rupture, and asymptomatic PAA without symptoms or limb threatening events. Patient and anatomic factors based on duplex ultrasound (DUS) were evaluated as potential predictors of symptom groups and thresholds of anatomic variables were identified using receiver operating characteristic curves.

**Results:** There were 470 PAAs identified in 331 patients. The mean age was 74 years at diagnosis, 94% of patients were white, and 97% of patients were male. Cardiovascular comorbidities were prevalent in and similar between all patient groups studied, and almost all patients were on anticoagulation or antiplatelet therapy at the time of diagnosis, at 96%. The most common concurrent aneurysm was abdominal aortic aneurysm (62%). Patient comorbidities were not associated with symptom status. PAAs with a higher percent thrombus burden were 18 times more likely to experience a limb threatening event and 24 times more likely to experience a TE. A largest diameter threshold of 20.4 mm was predictive of a TE (sensitivity 78.1%, specificity 40.5%), but percent thrombus threshold of 62% outperformed largest diameter as a predictor of a TE (sensitivity of 81.3%, specificity 52.0%). Percent thrombus threshold of 73% also predicted limb threatening events (sensitivity 65.7%, specificity 69.7%).

**Conclusion:** A PAA diameter greater than 20.4 mm was predictive of TEs, which is in agreement with clinical practice guidelines. However, percent thrombus greater than 62% outperformed largest diameter as a predictor of a TE. This analysis supports the use of size greater than 20.4 mm and 62% thrombus in identifying high risk PAAs that warrant repair.

## INTRODUCTION

Popliteal artery aneurysms (PAAs) are the most common peripheral arterial aneurysm, accounting for 70% of all peripheral aneurysms^1^, with an incidence of 10.4 cases per million inhabitants per year^2,3^. PAAs most commonly occur in 60- to 70-year-old men and are associated with presence of an abdominal aortic aneurysms (AAA)^4^. The most serious complication of typical aneurysmal disease is rupture, but rupture of PAAs is rare^5^. PAAs do not have the typical aneurysmal disease complication of rupture because they are contained within the confined space of the popliteal fossa. The most common serious complication of a PAA is instead occurrence of thrombosis or thromboembolic events (TEs) resulting in acute limb ischemia^5^. Acute presentations require unplanned interventions, which are associated with higher rates of earlier amputation and death than elective surgical repair^6^. Definitive surgical repair options include the gold standard open surgical bypass with a venous conduit^3,7^ or minimally invasive endovascular repair with covered stent placement^8^. While there is current debate surrounding which repair strategy is best, both open and endovascular repair of PAAs pose a risk of wound complications, need for additional procedures, and amputation ^2,3,6,9^.

PAAs are associated with high limb amputation rates and mortality^2^, but operative repair also poses risk to patients. Therefore, the Society for Vascular Surgery (SVS) has published a set of guidelines for repair of high risk PAAs^9^. Currently, the grade 1 recommendation for a PAA with a diameter greater than 20 mm on duplex ultrasound (DUS) is surgical repair. However, there is only moderate quality of evidence for this guideline given the debate surrounding the relationship between PAA size and associated complications^9^. There is one study of 106 patients in 1994 that specifically suggests a cutoff of 20 mm for identifying high risk PAAs^10^. Other studies have found larger sizes are associated with complications: acutely thrombosed PAAs have an aneurysm diameter larger than 25 mm^6,11^ and ruptured PAAs have even still a larger diameter compared to unruptured PAAs^5^.

Some studies show size does not confer increased risk of TEs or need for emergent surgery^12^. A study conducted in 2021 of 397 aneurysms in Sweden and New Zealand found there was no association between the size of the PAA and the need for emergent surgery due to clinical symptoms^13^. Another study showed that smaller PAAs were associated with higher incidence of thrombosis and clinical symptoms compared to larger PAAs^14^. The paucity of data surrounding PAA characteristics associated with adverse events has led some institutions to recommend repair of any PAA, regardless of size or symptoms^15^. Large studies of over 300 PAAs in Germany^11^ and the Netherlands^1^ have not identified high-risk features of PAAs and advocate for prophylactic repair of all PAAs on identification.

Efforts to identify PAA characteristics associated with PAA symptoms or adverse events have been limited to clinical variables and size. The natural history studies available are from the 1990’s^1,15^ or conducted in very small cohorts of patients^15^. Large contemporary studies are focused on comparing endovascular and open methods^6^, perioperative outcomes^2^, and long-term patency rates^6,11^. There has only been one study to our knowledge that attempted to identify a cutoff size associated with increased risk of thrombosis based on a threshold of 30-mm diameter and greater than 45 degrees of distortion, which is not a simple measurement that can be performed on routine DUS^16^. In this study, we specifically focus on identifying thresholds for PAA measurements on DUS associated with TE and limb threatening events specifically from PAA, including acute limb ischemia (ALI) and rupture. Identification of PAA measurements other than size associated with high risk of complications will risk stratify patients for surgery and guide criteria for repair.

## METHODS

### Cohort of Interest

The Mass General Brigham Human Research Committee Institutional Review Board approved the study protocol for patients >18 years of age and patient consent to participate was waived (IRB # 2019P003163). The cohort was derived from the Massachusetts General Brigham (MGB) Research Patient Data Registry (RPDR), a multi-institutional repository that gathers demographics, diagnosis codes, encounter data, procedural codes, medications, and other patient clinical information. The RPDR database was queried for all patients with a diagnosis code for “Aneurysm of the artery of lower extremity” (ICD9 442.3/ ICD10 I72.4) from the inception of the database in 2008 to 2022, which resulted in 3,503 patients. Manual chart review of each of the 3,503 patients by authors T.R.B. and B.G. was then performed to confirm the presence of a PAA. PAA was defined as greater than 1cm on any imaging modality, as the standard definition of an aneurysm is a diameter greater than 1.5 times the normal size of the artery^17^. If there was a discrepancy, senior author A.D. made a final determination. Baseline covariates were obtained by manual chart review for each patient at the time of diagnosis. Covariates collected included age at diagnoses of PAA, self-reported race, self-reported ethnicity, anticoagulation status, smoking status, cardiovascular comorbidities, and other reported aneurysms. Patients were also reviewed and excluded if a connective tissue disorder was present.

Symptom status at the time of intervention or at the time of last documentation was determined through review of clinical documentation including admission notes, consultation notes, clinic visit notes, operative reports, and discharge summaries. Chronic Limb Ischemia (CLI) was defined as chronic ischemic rest pain, ulceration or gangrene attributable to the occlusion of peripheral arterial vessels. ALI was defined as a rapid decrease in lower limb blood flow due to acute occlusion of peripheral artery. TE was defined as thrombosis of vessels distal to the PAA originating from the popliteal thrombus burden. An imaging modality of DUS, Computed Tomography Angiography (CTA), or lower extremity angiogram was reviewed for documented occlusion of a distal vessel or the PAA itself. All TEs documented as an outcome in this study occurred before any operative repair occurred. Symptomatic PAA was defined as documented claudication or chronic limb ischemia (CLI). Limb threatening PAA was defined as a PAA with a TE, ALI, or rupture of the PAA attributed to the presence of a PAA. Asymptomatic PAA was defined as the absence of symptoms or limb threatening events as described above.

All operative details were obtained from operative reports or vascular surgeon documented post-operative visits. The primary indication for an intervention was defined as the indication listed as reason for surgery in the operative report or post-operative note. Emergent intervention was defined as listing the case as an emergent operation in the operative report. Amputation was not counted as a primary intervention, as amputation secondary to PAA related events was an outcome measure. Endovascular intervention was defined as if either the primary component or a secondary component of the initial operation was performed endovascularly. Return to the operating room (OR) was defined as return to the operating room within 72 hours to correct a procedural related complication. Death was recorded as a date of deceased within 5 years of PAA diagnosis.

DUS characteristics of PAAs were abstracted by RPVI certified physician for noninvasive vascular interpretation (G.G.). G.G. was blinded to ultrasound reports and annotated all DUS for all characteristics that may be important for a TE. The definition of each abstracted characteristic is listed below. Surface transversal area was defined as the surface of the popliteal aneurysm in cross section, with inclusion of the arterial wall, from one DUS still image. Internal surface area was defined as the surface of the popliteal aneurysm in cross section, without inclusion of the arterial wall, from one DUS still image. Patent channel area was defined as surface of the circulating arterial lumen, free from thrombus, calculated from one DUS still image. Thrombus area was defined as the area within the artery occupied by thrombus material, calculated by subtracting the area of the true lumen from the area of the entire aneurysm (Figure 1). Anteroposterior diameter was defined as the largest diameter in any still image measured in the anterior to posterior dimension. Transverse diameter was defined as the largest diameter in any still image measured in the transverse dimension. Largest diameter was defined as the maximum diameter annotated in either anteroposterior or transverse dimensions. Percent thrombus was calculated by dividing the thrombus area by the area of the entire aneurysm by multiplied by 100 (Figure 1).

**Figure 1.**
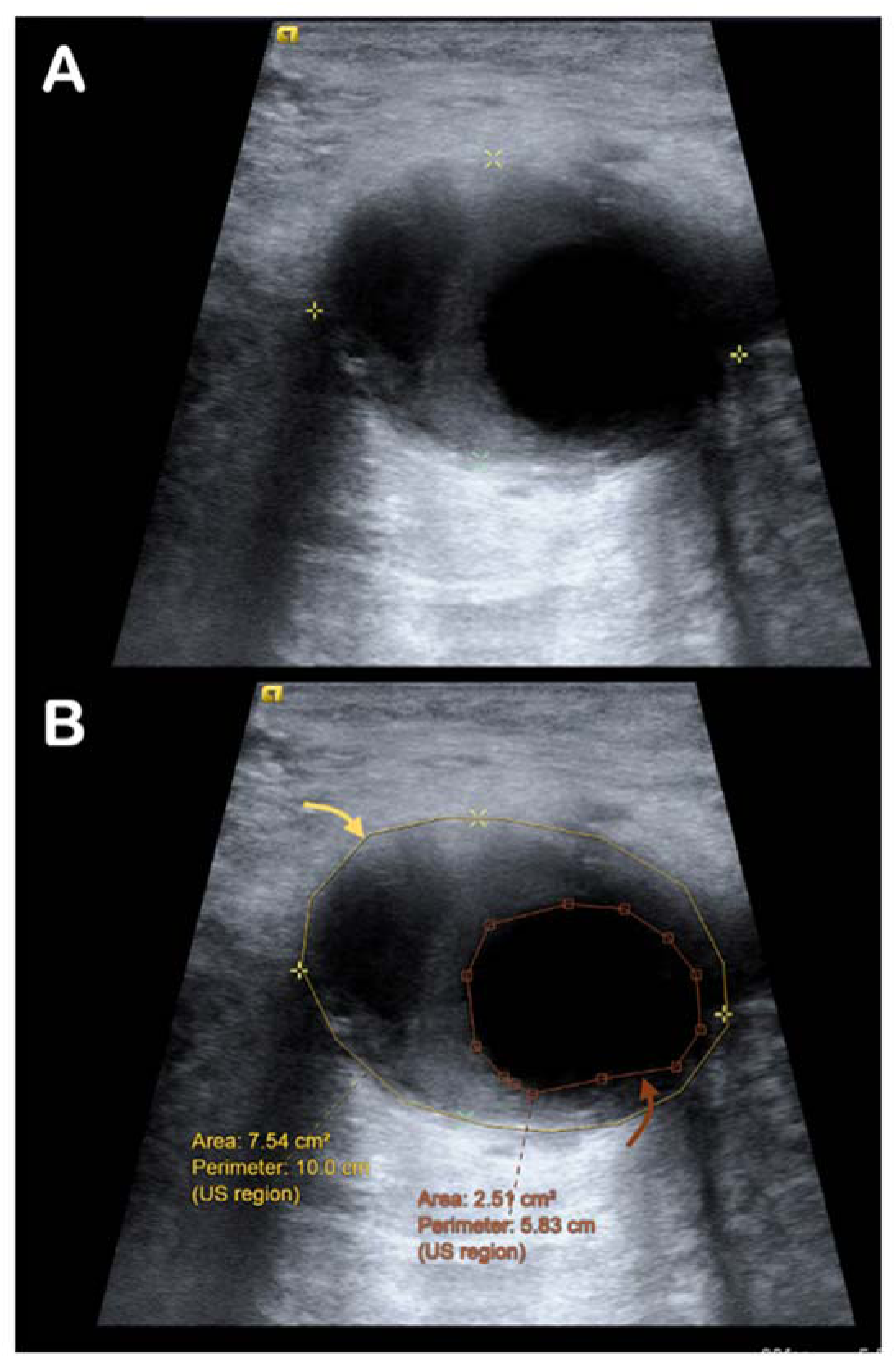
Duplex ultrasound measurements of popliteal artery aneurysm percent thrombus. (A) The B-mode image with the largest diameter in the transverse section of the aneurysm was selected for analysis. (B) Measurements taken included the entire area of the aneurysm and the area of the true lumen: the yellow ellipse contouring the outer wall of the aneurysm was used to calculate the area of the entire aneurysm. The orange ellipse contouring the arterial true lumen was used to calculate the area of the true lumen. The thrombus area was calculated by subtracting the area of the true lumen from the area of the entire aneurysm. Percent thrombus was calculated by dividing the thrombus area by the area of the entire aneurysm by multiplied by 100.

### Statistical Analysis

Patient demographics were expressed for PAAs based on symptom status using descriptive statistics in Table 1. Each PAA was treated as a separate event. Measurements from PAA DUS were expressed in terms of median and IQR given the non-parametric distribution of data. Univariate analysis was conducted using multinomial logistic regression models. Subsequent multinomial logistic regression models included age as a covariate. Models that also included covariates of race, sex, and/or anticoagulation produced unstable estimates due to uneven distributions of the data. Thresholds identified for PAA DUS measurements in relation to PAA symptom status were identified using receiver operating characteristic curves (ROCs), where the decision point on the ROC curve closest to achieving a sensitivity of 1 and a specificity of 1 were selected. These thresholds were then graphically represented in Kaplan-Meier curves generated for outcomes. Statistical analyses were performed using STATA, version 15.1 (StataCorp, College Station, Texas). P-value of <0.05 was considered statistically significant for all analyses.

**Table 1.** Characteristics and Demographics. 331 patients with 470 popliteal artery aneurysms and their demographic characteristics separated by symptoms on presentation. P-values calculated across symptom presentation were calculated by Fisher’s Exact test.

| Demographics<br>n (%) | Asymptomatic<br>(n=230) | Symptomatic<br>(n=127) | Limb<br>Threatening<br>(n=113) | Total<br>(n=470) | P-value |
| --- | --- | --- | --- | --- | --- |
| Age, median (IQR) | 74 (66, 81) | 78 (72, 62) | 73 (62, 78) | 73 (65, 79) | 0.02* |
| Race (white) | 222 (97%) | 102 (90%) | 118 (93%) | 442 (94%) | 0.06 |
| Sex (male) | 225 (98%) | 110 (97%) | 121 (95%) | 456 (97%) | 0.38 |
| Laterality |  |  |  |  | 0.93 |
| Left | 113 (49%) | 53 (47%) | 61 (48%) | 227 (48%) |  |
| Right | 117 (51%) | 60 (53%) | 66 (52%) | 243 (52%) |  |
| Ever smoker | 155 (67%) | 90 (80%) | 92 (72%) | 337 (72%) | 0.02* |
| Hypertension | 210 (91%) | 96 (85%) | 116 (91%) | 422 (90%) | 0.12 |
| Hyperlipidemia | 188 (82%) | 91 (81%) | 109 (86%) | 388 (83%) | 0.50 |
| Coronary artery<br>disease | 112 (49%) | 63 (56%) | 66 (52%) | 241 (51%) | 0.46 |
| Type 1 diabetes | 1 (0%) | 0 (0%) | 2 (2%) | 3 (1%) | 0.32 |
| Type 2 diabetes | 69 (30%) | 37 (33%) | 36 (28%) | 142 (30%) | 0.76 |
| Dialysis | 20 (9%) | 6 (5%) | 9 (7%) | 35 (7%) | 0.52 |
| Atrial fibrillation | 75 (33%) | 39 (35%) | 42 (33%) | 156 (33%) | 0.94 |
| DVT | 62 (27%) | 36 (32%) | 41 (32%) | 139 (30%) | 0.48 |
| Anticoagulation | 223 (97%) | 110 (97%) | 118 (93%) | 451 (96%) | 0.15 |
| Aspirin | 209 (91%) | 103 (91%) | 110 (87%) | 422 (90%) | 0.38 |
| Plavix | 86 (37%) | 55 (49%) | 57 (45%) | 198 (42%) | 0.11 |
| DOAC | 52 (23%) | 16 (14%) | 14 (11%) | 82 (17%) | 0.01* |
| Warfarin | 122 (53%) | 56 (50%) | 58 (46%) | 236 (50%) | 0.41 |
| enoxaparin | 81 (35%) | 38 (34%) | 44 (35%) | 163 (35%) | 0.96 |
| Anuerysm |  |  |  |  |  |
| Abdominal aorta | 157 (68%) | 63 (56%) | 80 (63%) | 300 (64%) | 0.08 |
| Descending aorta | 49 (21%) | 19 (17%) | 15 (12%) | 83 (18%) | 0.07 |
| Ascending aorta | 22 (10%) | 4 (4%) | 14 (11%) | 40 (9%) | 0.07 |
| Carotid artery | 4 (2%) | 0 (0%) | 3 (2%) | 7 (1%) | 0.32 |
| Iliac artery | 94 (41%) | 39 (35%) | 45 (35%) | 178 (38%) | 0.42 |
| Other | 70 (30%) | 41 (36%) | 39 (31%) | 150 (32%) | 0.52 |

## RESULTS

### Demographics

A total of 331 patients who had 470 PAAs were included in this analysis. These patients were on average 74 years old at age of diagnosis, 94% of patients were white, and 97% of patients were male (Table 1). These demographics were not significantly different between the asymptomatic, symptomatic, and TE groups. PAAs were equally identified in both the left side at 48% and the right side at 52%. The occurrence of ever smoking was significantly higher in both symptomatic PAAs (80%) and limb threatening PAAs (72%) compared to asymptomatic PAAs (67%). Cardiovascular comorbidities were prevalent in and similar between all patient groups studied, including hypertension (90%), hyperlipidemia (83%), coronary artery disease (51%), and atrial fibrillation (33%) (Table 1). Type 2 diabetes was more prevalent at 30% than type I diabetes at 1%. Severe kidney disease requiring dialysis was present in 7% of patients. Almost all patients were on anticoagulation or antiplatelet therapy at the time of diagnoses (96%), with aspirin in 90% of patients, plavix in 42% of patients, DOAC in 17% of patients, warfarin in 50% of patients, and enoxaparin in 35% of patients (Table 1). The most common concurrent aneurysm was abdominal aortic aneurysm (62%). Other aneurysms were less common, with descending aortic aneurysms in 18%, ascending aortic aneurysms in 9%, carotid artery aneurysms in 1%, iliac artery aneurysms in 38%, and other aneurysms in 32% of patients. There was a family history of aneurysms in 4% of patients.

### Popliteal Aneurysm Clinical and Operative Characteristics

At the time of initial diagnoses, patients with popliteal artery aneurysms presented with a variety of symptoms: the size of the aneurysm was measured greater than 2cm in 75%, mural thrombus was present in 54%, claudication affected 41%, chronic limb ischemia (CLI) was present in 6%, acute limb ischemia (ALI) was present in 14%, the aneurysm was occluded in 22%, the aneurysm was ruptured in 1%, and a TE was detected in 12% (Table 2). There were 58 PAAs that had a TE: 62% occurred in arteries below the knee, 62% occurred in arteries above the knee but below the PAA, 48% had petechia, and 47% occurred in toe vessels. Regardless of symptom presentation, the majority of patients with a PAA overall underwent intervention at 68%: asymptomatic PAAs underwent intervention at 53%, which was significantly less than symptomatic PAAs at 69% and PAAs with a limb threatening event at 95% (Table 3). PAAs with a limb threatening event underwent the greatest proportion of emergent surgeries at 59%, followed by symptomatic PAAs at 2%, and lastly asymptomatic PAAs at 2%. The most common indication for intervention overall was size greater than 20 mm. The most common indication for intervention, by subgroup, was size greater than 20 mm at 64% in asymptomatic PAAs, but mural thrombus for symptomatic PAAs at 42% and ALI for PAAs with a limb threatening event at 46%. Of PAAs that underwent open intervention, the medial incision was more commonly made than the posterior incision across all symptom groups at 75%. The most common conduit used in open intervention across all symptom groups was autologous vein for a bypass at 67%, and the most common interposition conduit used was synthetic graft for 38 PAAs compared to vein for 3 PAAs. Initial endovascular intervention was performed significantly more frequently in symptomatic PAAs at 22% and PAAs with a limb threatening event at 32% compared to asymptomatic PAAs at 17%. Of PAAs that underwent endovascular intervention, the most common intervention in both asymptomatic and symptomatic PAAs was Viabahn stent placement at 95% and 74% respectively. The most common endovascular intervention in PAAs with a limb threatening event was thrombolysis at 41% followed by Viabahn stent placement at 39%. There were 11 PAAs that required a return to the operating room (OR) most commonly for acute thrombosis in 5 PAAs without any significant differences among symptom groups.

**Table 2.**
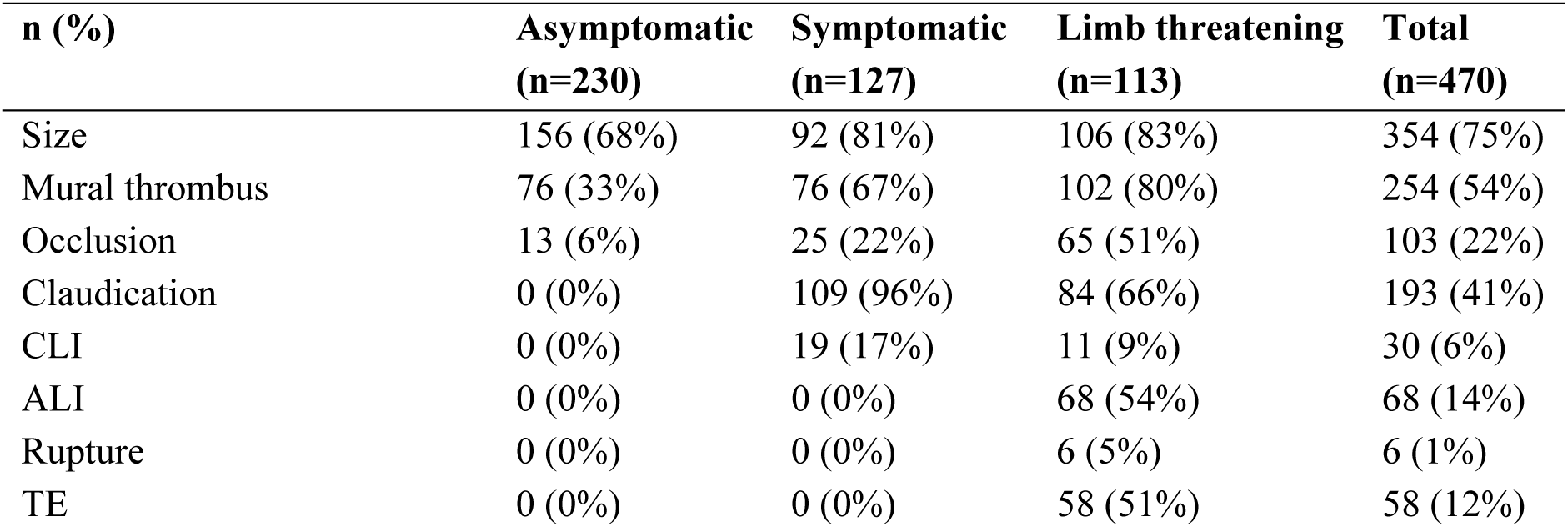
Symptoms of popliteal artery aneurysms on presentation. 470 popliteal artery aneurysms among 331 patients and relevant symptoms on presentation. Abbreviations: CLI, chronic limb ischemia; ALI, acute limb ischemia; TE, thromboembolism.

**Table 3.** Characteristics of initial intervention for popliteal artery aneurysms. Details for intervention performed on 318 popliteal artery aneurysms derived directly from operative reports. P-values calculated across symptom presentation were calculated by Fisher’s Exact test. Abbreviations: TE, thromboembolism; CLI, chronic limb ischemia; ALI, acute limb ischemia; OR, operating room; PTA, percutaneous transluminal angioplasty.

| n/ total n (%) | Asymptomatic | Symptomatic | Limb threatening | Total | P-value* |
| --- | --- | --- | --- | --- | --- |
| **Intervention** | 123/230 (53%) | 88/127 (69%) | 107/113 (95%) | 318/470 (68%) | <0.01* |
| **Emergent** | 2/123 (2%) | 2/88 (2%) | 63/107 (59%) | 67/318 (22%) | <0.01* |
| **Primary Indication** |  |  |  |  | <0.01* |
| Size | 74/123 (64%) | 24/88 (28%) | 4/107 (4%) | 102/318 (32%) |  |
| Mural thrombus | 34/123 (29%) | 36/88 (42%) | 19/107 (18%) | 89/318 (29%) |  |
| Occlusion | 5/123 (4%) | 7/88 (8%) | 21/107 (20%) | 33/318 (11%) |  |
| Claudication | 1/123 (1%) | 4/88 (5%) | 2/107 (2%) | 7/318 (2%) |  |
| CLI | 0/123 (0%) | 13/88 (15%) | 7/107 (7%) | 20/318 (6%) |  |
| ALI | 0/123 (0%) | 0/88 (0%) | 50/107 (46%) | 50/318 (16%) |  |
| Rupture | 0/123 (0%) | 0/88 (0%) | 5/107 (5%) | 5/318 (2%) |  |
| Other | 1/123 (1%) | 2/88 (2%) | 0/107 (0%) | 3/318 (1%) |  |
| **Open Incision** | 97/123 (79%) | 70/88 (80%) | 83/107 (78%) | 250/318 (79%) | 0.55 |
| Posterior | 26/97 (27%) | 20/70 (29%) | 16/83 (19%) | 62/250 (25%) |  |
| Medial | 71/97 (73%) | 50/70 (71%) | 67/83 (81%) | 188/250 (75%) |  |
| **Conduit for Bypass** | 97/123 (79%) | 70/88 (80%) | 82/107 (77%) | 249/318 (78%) | 0.56 |
| Autologous Vein | 66/97 (68%) | 42/70 (60%) | 58/82 (71%) | 166/249 (67%) |  |
| Cadaver Vein | 0/97 (0%) | 1/70 (1.4%) | 0/82 (0%) | 1/249 (0%) |  |
| Graft | 16/97 (16%) | 11/70 (16%) | 14/82 (17%) | 41/249 (16%) |  |
| Interposition Graft | 15/97 (15%) | 15/70 (21%) | 8/82 (10%) | 38/249 (15%) |  |
| Interposition Vein | 0/97 (0%) | 1/70 (1%) | 2/82 (2%) | 3/249 (1%) |  |
| **Endovascular** | 21/123 (17%) | 19/88 (22%) | 34/107 (32%) | 74/318 (23%) | 0.03* |
| Angiogram | 1/21 (5%) | 3/19 (16%) | 3/34 (9%) | 7/74 (9%) |  |
| PTA | 0/21 (0%) | 0/19 (0%) | 1/34 (3%) | 1/74 (1%) |  |
| Viabahn Stent | 20/21 (95%) | 14/19 (74%) | 10/34 (29%) | 44/74 (59%) |  |
| Thrombolysis | 0/21 (0%) | 1/19 (5%) | 14/34 (41%) | 15/74 (20%) |  |
| Thrombectomy | 0/21 (0%) | 1/19 (5%) | 6/34 (18%) | 7/74 (9%) |  |
| **Return to OR** | 2/123 (2%) | 1/88 (1%) | 8/107 (7%) | 11/318 (3%) | 0.13 |
| Acute thrombosis | 1/2 (50%) | 0/1 (0%) | 4/8 (50%) | 5/11 (45%) |  |
| Compartment syndrome | 0/2 (0%) | 0/1 (0%) | 2/8 (25%) | 2/11 (18%) |  |
| Hematoma | 1/2 (50%) | 0/1 (0%) | 0/8 (0%) | 1/11 (9%) |  |
| Seroma | 0/2 (0%) | 1/1 (100%) | 0/8 (0%) | 1/11 (9%) |  |
| Stenosis | 0/2 (0%) | 0/1 (0%) | 1/8 (13%) | 1/11 (9%) |  |
| Other | 0/2 (0%) | 0/1 (0%) | 1/8 (13%) | 1/11 (9%) |  |

### Popliteal Aneurysm Ultrasound Characteristics

The pre-operative DUS performed closest to the time of operation or the most recent DUS was used to calculate characteristics of each PAA (Table 4). Aneurysm diameters calculated varied by measurement type. The largest diameter was significantly smaller for asymptomatic PAAs at 22.1 mm (IQR 16.5-31.1) compared to symptomatic PAAs at 23.2 mm (IQR 19.0-29.0) and PAAs with a limb threatening event at 27.7 mm (IQR 20.5-37.9) P-value = 0.02. Median anteroposterior diameter was not significantly different between asymptomatic PAAs at 20.5 mm (IQR 16.2-27), symptomatic PAAs at 21.5 mm (IQR 18.4-27.4), and PAAs with a limb threatening event at 24.7 mm (IQR 18.4-33.9). The median transverse diameter of asymptomatic PAAs at 21.3 mm (IQR 16-29.2) was significantly smaller than both symptomatic PAAs at 21.8 mm (IQR 17.2-30.3) and PAAs with a limb threatening event at 27.6 mm (IQR 20.6, 35.2) P-value = 0.02. Total surface areas were also calculated, with significantly smaller median total surface area in asymptomatic PAAs at 339 mm^2^ (IQR 205-596.5) compared to symptomatic PAAs at 399 mm^2^ (IQR 266-623) and PAAs with a limb threatening event at 602 mm^2^ (308-940) P-value = 0.02. Median patent channel area was the largest in asymptomatic PAAs at 152 mm^2^ (IQR 90.6-214) compared to symptomatic PAAs at 83.6 mm^2^ (IQR 44.2-202) and PAAs with a limb threatening event at 128 mm^2^ (IQR 44.3-215) P-value = 0.03. Median thrombus area was the smallest in asymptomatic PAAs at 157 mm^2^ (IQR 21-395) compared to symptomatic PAAs at 250 mm^2^ (IQR 119-461) and PAAs with a limb threatening event at 388 mm^2^ (190-770) P-value = 0.01. Median patent channel area and thrombus area were used to calculate a percent thrombus in all 470 aneurysms in 331 patients, which was significantly larger in PAAs with a limb threatening event at 80% and symptomatic PAAs at 80% compared to asymptomatic PAAs at 50% P-value < 0.01.

**Table 4.** Characteristics of popliteal artery aneurysms on duplex ultrasound. The characteristics of 470 popliteal artery aneurysms measured directly from duplex ultrasound (DUS) images. All measurements calculated were derived from the pre-operative DUS performed closest to the time of the operation or the most recent DUS documented. P values calculated across symptom presentation were calculated by Kruskal Wallis test for linear measurements and Fisher’s Exact test for categorical measurements. Abbreviations: TE, thromboembolism; AP, Anterior Posterior.

| **Median (IQR)** | **Asymptomatic (n=230)** | **Symptomatic (n=172)** | **Limb threatening (n=113)** | **Total (n=470)** | **P-value** |
| --- | --- | --- | --- | --- | --- |
| Largest diameter (mm) | 22.1 (16.5, 31.1) | 23.2 (19.0, 29.0) | 27.7 (20.5, 37.9) | 22.8 (17.7, 31.9) | 0.02* |
| AP diameter (mm) | 20.5 (16.2, 27.0) | 21.5 (18.4, 27.4) | 24.7 (18.4, 33.9) | 21.3 (16.8, 28.5) | 0.08 |
| Transverse diameter | 21.3 (16, 29.2) | 21.8 (17.2, 30.3) | 27.6 (20.6, 35.2) | 22.1 (16.6, 30.4) | 0.02* |
| Total surface (mm<sup>2</sup>) | 339 (205, 596.5) | 399 (266, 623) | 602 (308, 940) | 382 (230, 742) | 0.02* |
| Patent channel area (mm<sup>2</sup>) | 152 (90.6, 214) | 83.6 (44.2, 202) | 128 (44.3, 215) | 133.5 (75, 213) | 0.03* |
| Thrombus area (mm<sup>2</sup>) | 157 (21, 395) | 250 (119, 461) | 388 (190, 770) | 234 (72, 479) | 0.01* |
| Percent thrombus (%) | 50 (10, 80) | 80 (60, 90) | 80 (50, 90) | 70 (30, 80) | <0.01* |

### Popliteal Aneurysm Regression Analysis

Models were created to assess the association of PAA characteristics with PAA symptom status. Given TE was a limb threatening event of particular interest, PAAs with TEs were evaluated as a separate outcome and relative risk ratios (RRR) were calculated. There were no significant univariate associations between all demographic factors listed in Table 1 and symptomatic PAA, PAA with a TE, and adverse events. PAA DUS measurements listed in Table 4 were tested for a univariate association with symptom status in Table 5. Higher percent thrombus was significantly associated with symptomatic PAAs (RRR 16.2; CI 2.4-109.8; p<0.01), PAAs with a limb threatening event (RRR 11.8; CI 1.8-76.7; p=0.01), and PAAs with a TE (RRR 13.2; CI 4.1-42.6; p<0.01). Larger thrombus area was significantly associated with limb threatening PAA (OR 1.01; CI 1.00-1.02; p=0.03), but not symptomatic PAAs (RRR 1.01; CI 1.00-1.02; p=0.16) or PAA with a TE (RRR 1.00; CI 0.97-1.03; p=0.81) (Table 5). Largest diameter, channel area, and mural thrombus did not have any association with symptom status. Given nearly all patients were white males on anticoagulation therapy, multivariate models were created using age (Table 6). Higher percent thrombus was still significantly associated with PAA with a TE (RRR 24.3; CI 1.44-410; p=0.03), PAAs with a limb threatening event (RRR 17.9; CI 3.76-85.0; p<0.01), and symptomatic PAAs (RRR 15.2; CI 2.69-72.3; p<0.01). After including age, channel area, thrombus area, largest diameter, and mural thrombus did not have any significant association with symptom status or limb threatening events.

**Table 5.** Popliteal artery aneurysm (PAA) characteristics associated with symptomatic PAA, limb threatening PAA, or PAA with thromboembolic (TE) events in a univariate model. All diameters were calculated in cm and areas in cm^2^.

|  | **RRR** | **95% CI**<br>**Lower** | **95% CI**<br>**Upper** | **P-Value** |
| --- | --- | --- | --- | --- |
| **Percent thrombus** |  |  |  |  |
| Symptomatic | 16.2 | 2.4 | 109.8 | <0.01* |
| Limb threatening | 11.8 | 1.8 | 76.7 | 0.01* |
| TE | 13.2 | 4.1 | 42.6 | <0.01* |
| **Largest diameter** |  |  |  |  |
| Symptomatic | 1.04 | 0.97 | 1.12 | 0.25 |
| Limb threatening | 0.97 | 0.97 | 1.18 | 0.75 |
| TE | 1.00 | 0.97 | 1.03 | 0.81 |
| **Channel area** |  |  |  |  |
| Symptomatic | 0.99 | 0.97 | 1.02 | 0.66 |
| Limb threatening | 0.97 | 0.93 | 1.01 | 0.12 |
| TE | 0.99 | 0.99 | 1.00 | 0.07 |
| **Thrombus area** |  |  |  |  |
| Symptomatic | 1.01 | 1.00 | 1.02 | 0.16 |
| Limb threatening | 1.01 | 1.00 | 1.02 | 0.03* |
| TE | 1.00 | 0.97 | 1.03 | 0.81 |
| **Mural Thrombus** |  |  |  |  |
| Symptomatic | NA | NA | NA | NA |
| Limb threatening | 2.25 | 0.46 | 11.6 | 0.32 |
| TE | NA | NA | NA | NA |

**Table 6.**
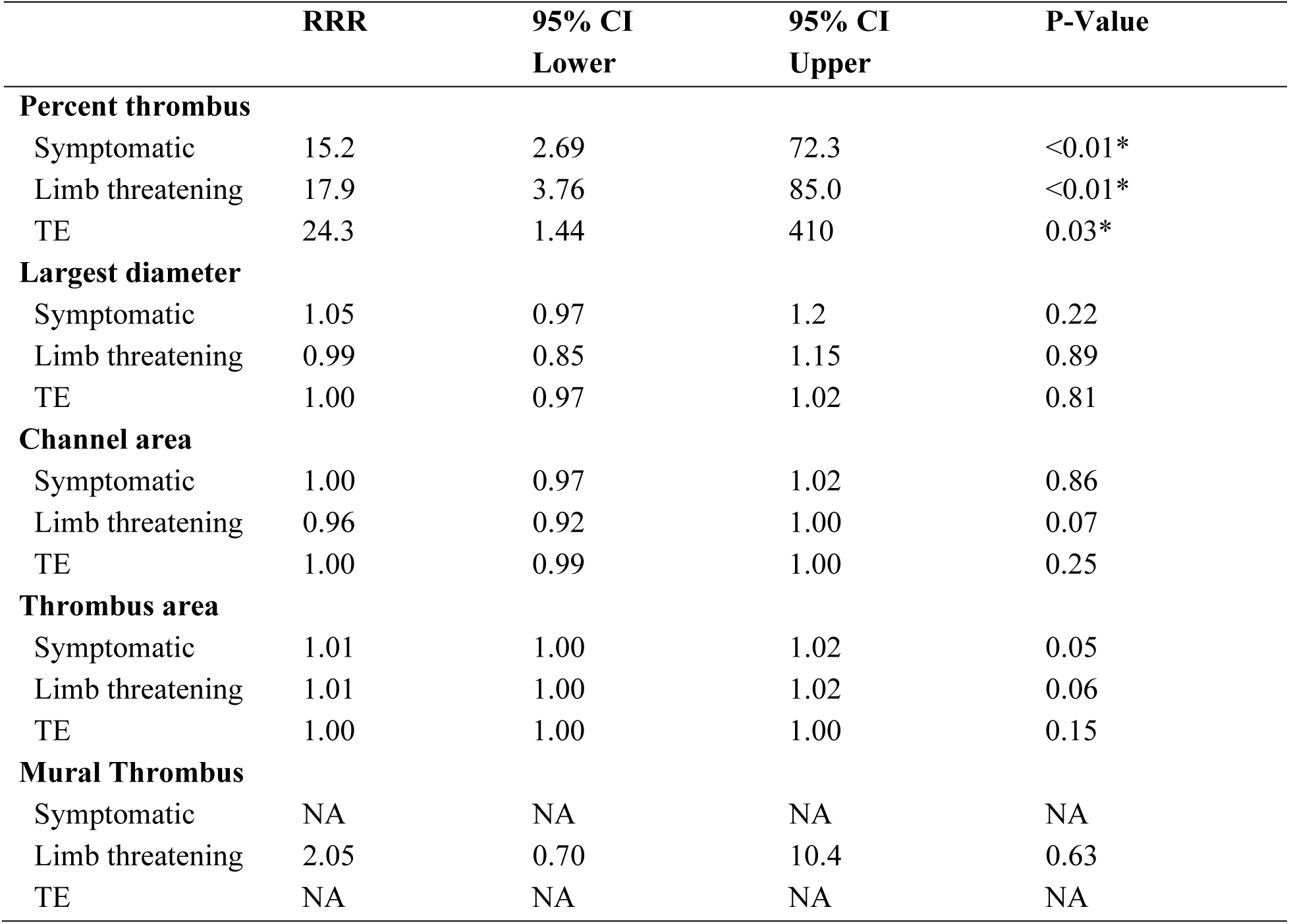
Popliteal artery aneurysms (PAA) characteristics associated with symptomatic PAA, limb threatening PAA, or PAA with TE in a multivariable model controlling for age.

Using these data, cut points were identified for TEs and limb threatening events in Table 7. For TEs, percent thrombus at a threshold of 62% had the highest specificity at 52% and a good sensitivity of 81.3%. Largest diameter at 20.4 mm had the lowest sensitivity at 78.1% and a specificity of 40.5%. For limb threatening events, thrombus area at a threshold of 114 mm^2^ had the highest sensitivity at 77.4% and largest diameter at a threshold of 27.9 mm had the highest specificity at 65.7%. Kaplan Meier curves in Figure 2 demonstrate the ability of thresholds on PAA DUS to predict a TE, including a percent thrombus of 62%, a largest diameter of 20.4 cm, a channel area of 114 mm^2^, and a thrombus area of 165 mm^2^. Figure 3 demonstrates the ability of thresholds on PAA DUS to predict limb threatening events, including percent thrombus threshold of 73%, largest diameter threshold of 27.9 mm, channel area threshold of 114 mm^2^, and a thrombus area threshold of 114 mm^2^.

**Figure 2.**
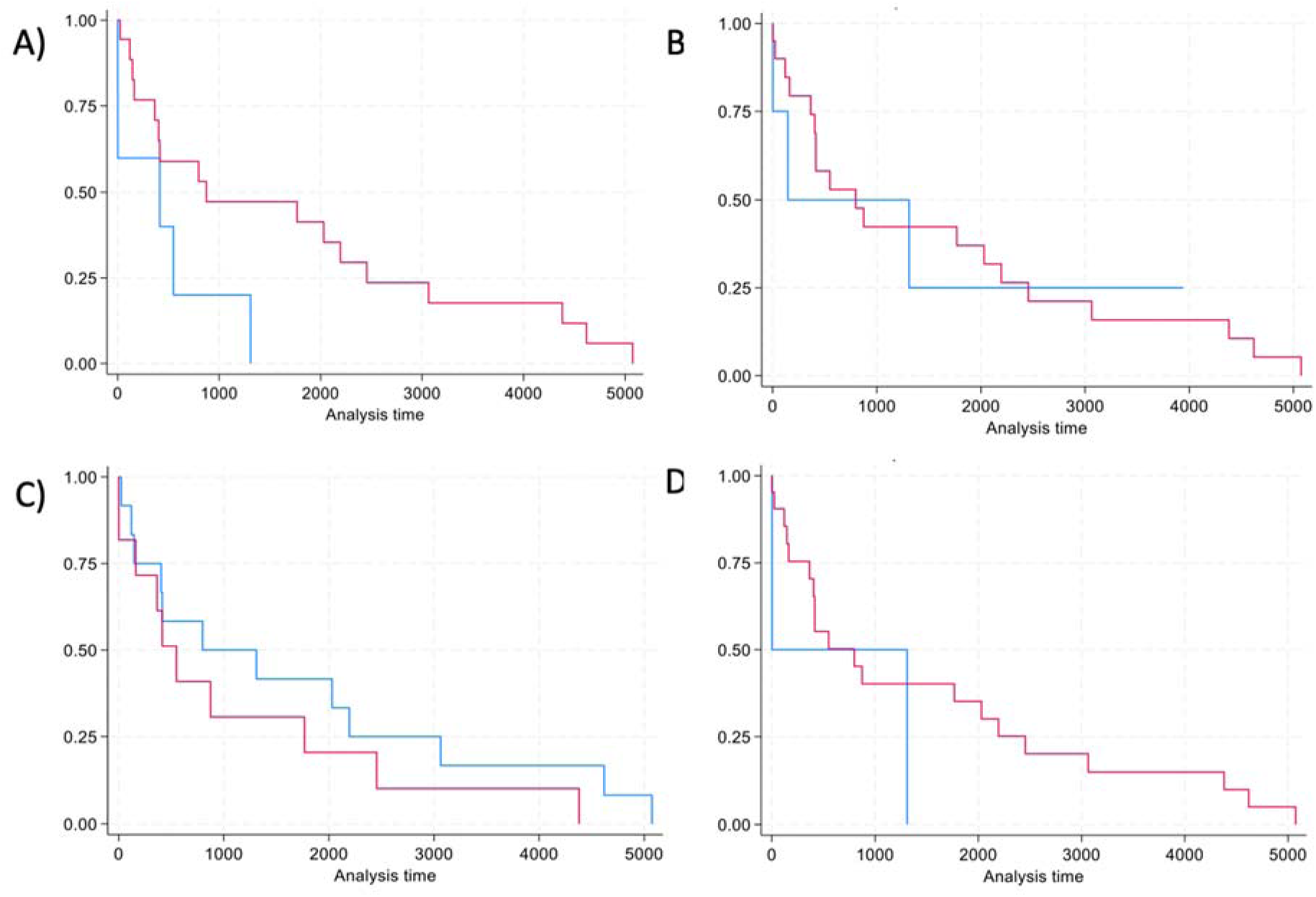
Kaplan Meier curves showing time to Thromboembolic Event (TE) stratified by ultrasound characteristic of the popliteal artery aneurysm (PAA). The blue line represents PAAs below the threshold and the red line represents all PAAs above the threshold. A) Percent thrombus threshold of 62% B) largest diameter threshold of 20.4 mm, C) channel area threshold of 114 mm^2^, D) and a thrombus area threshold of 165 mm^2^.

**Figure 3.**
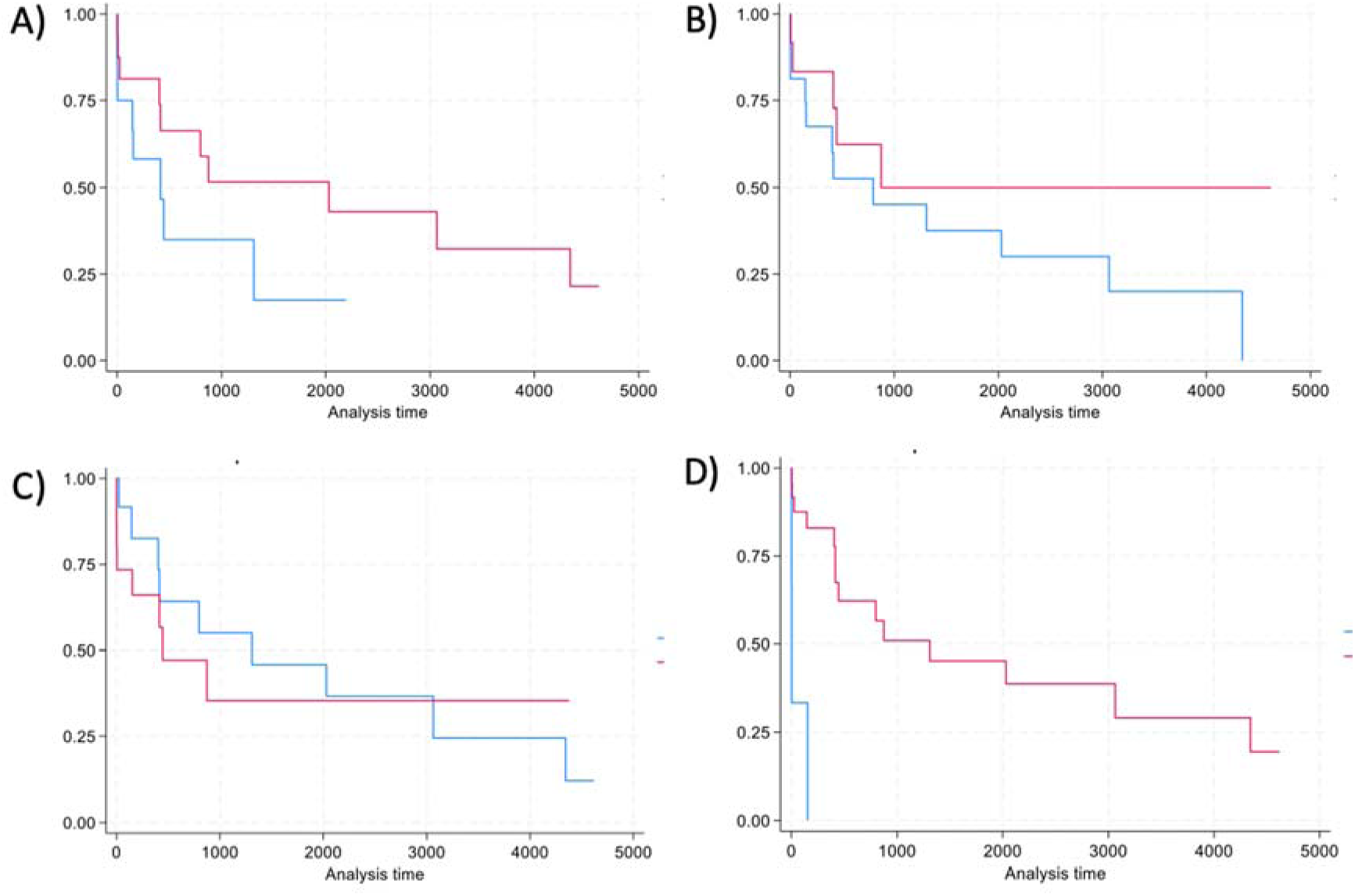
Kaplan Meier curves showing time to limb threatening event stratified by ultrasound characteristic of the popliteal artery aneurysm (PAA). Limb threatening event was defined as occurrence of a TE, ALI, rupture, or amputation. The blue line represents PAAs below the threshold and the red line represents all PAAs above the threshold. A) Percent thrombus threshold of 73% B) largest diameter threshold of 27.9 mm, C) channel area threshold of 114 mm^2^, D) and a thrombus area threshold of 114 mm^2^.

**Table 7.** Threshold values of popliteal artery aneurysm duplex ultrasound measurements for thromboembolic (TE) and limb threatening event outcomes.

| **TE** | **Cut point** | **Sensitivity** | **Specificity** | **PPV** | **NPV** |
| --- | --- | --- | --- | --- | --- |
| Percent thrombus (%) | 62.0 | 81.3 | 52.0 | 30.6 | 91.4 |
| Largest diameter (mm) | 20.4 | 78.1 | 40.5 | 25.0 | 87.9 |
| Channel area (mm<sup>2</sup>) | 114 | 35.3 | 36.1 | 12.4 | 68.8 |
| Thrombus area (mm<sup>2</sup>) | 165 | 83.9 | 47.1 | 28.9 | 91.9 |
| **Limb Threatening** | **Cut point** | **Sensitivity** | **Specificity** | **PPV** | **NPV** |
| Percent thrombus (%) | 73.0 | 69.7 | 62.2 | 67.6 | 88.8 |
| Largest diameter (mm) | 27.9 | 25.0 | 65.7 | 84.9 | 78.2 |
| Channel area (mm<sup>2</sup>) | 114 | 32.3 | 36.2 | 89.0 | 68.7 |
| Thrombus area (mm<sup>2</sup>) | 114 | 77.4 | 37.3 | 76.7 | 87.0 |

## DISCUSSION

Although SVS guidelines define high risk PAAs as having mural thrombus, poor run off, and a diameter greater than 2 cm, there is little evidence to support any of these suggestions^17^. We carefully curated a large cohort of 470 PAAs in 331 patients and manually annotated DUS images for various anatomic characteristics. In the largest American cohort of PAAs, we confirmed a cutoff of 20.4 mm was predictive of a TE (sensitivity 78.1%, specificity 40.5%) and a cutoff of 27.9mm was predictive of limb threatening events (sensitivity of 77.4%, specificity 65.7%). These data also show that a percent thrombus threshold of 62% outperformed size as a predictor of a TE (sensitivity of 81.3%, specificity 52.0%). The largest diameter of a PAA was not significantly associated with TEs or limb threatening events in a multivariate analysis. This analysis fills a knowledge gap given the significant practice differences nationally and internationally surrounding criteria of PAAs that indicate need for repair.

Traditional aneurysmal disease, including thoracic aortic aneurysms^18^, AAAs^19^, and visceral aneurysms^20^ have a significant risk of rupture and resulting mortality regardless of patient characteristics; therefore, studies have appropriately focused on identifying size criteria for prophylactic repair of these aneurysms^21–23^. However, PAAs are anatomically constrained by surrounding muscles and tendons of the leg^24^, making rupture a rare outcom^5^. The most common serious clinical outcomes of PAAs are thrombosis and TEs resulting in acute limb ischemia^5^ and can often be the first symptom on presentation^25^. In our study, nearly half of all PAAs were asymptomatic on presentation, while the presenting symptoms for the remaining PAAs were less common than reported in other studies. We found rupture in only 1% of our cohort whereas other studies have reported 2%^5^, 3%^5^, or even 9%^2^ rupture rates. ALI was more common than rupture on presentation in our cohort at 14%, consistent with other studies ranging from 21%^11^ to 60%^15^. There have been several approaches to study PAAs in the context of clinical presentation: more recent studies have compared asymptomatic PAAs to symptomatic PAAs^2^. The definition of symptomatic included a wide range of symptoms, including claudication, CLI, and even ALI. Although both claudication and ALI are technically symptoms, claudication is not considered limb threatening and does not require surgery. Therefore, we have analyzed these data in three different groups: symptomatic PAA was defined as documented claudication or chronic limb ischemia (CLI). Limb threatening PAA was defined as a PAA with a TE, ALI, or rupture of the PAA. Asymptomatic PAA was defined as the absence of symptoms or limb threatening events as described above.

This study of 331 patients with 470 PAAs were older at age of diagnosis^6^ but otherwise had representative demographics and comorbidities compared to other studies of natural history: most patients are white males and around half of patients also have an AAA^1,7,11^. All comorbidities recorded were not significantly different across symptom groups, consistent with other reports unable to establish an association between any clinical variables and PAA related adverse events^7,11^. Some operative characteristics did differ by symptom group. Current SVS guidelines recommend PAA repair for a diameter greater than 20 mm, which was the most common indication for repair of asymptomatic PAA in our study, or presence of mural thrombus, which was the most common indication for repair of a symptomatic PAA or PAA with a limb threatening event in our study^17^. The most common open intervention was an operative bypass using vein through a medial approach, which did not differ with symptom group and is consistent with existing literature^7^. Endovascular interventions were more common in PAAs with a limb threatening event, as the most common endovascular intervention for this group was thrombolysis. Overall, the most common endovascular intervention was placement of a Viabahn stent, which has also been reported in literature^11^. Return to OR within 72 hours also did not vary by symptom group. Given no association of demographic factors, comorbidities, or operative characteristics with symptom groups, we focused on identifying anatomic differences associated with symptom groups.

There are several measurable characteristics of general stenosis and aneurysms on DUS have been previously shown to associate with adverse outcomes. The presence or absence of mural thrombus specifically was identified as a significant predictor of sac expansion in PAAs^26,27^. Thrombus area and patent channel area has not been studied in the context of PAAs, but this has been studied in AAA: higher thrombus burden in the aortic wall of an AAA, even after repair, has been associated with solid organ infarction from possible emboli^28^. One study specific to PAAs was able to measure distortion and size which discriminated between symptomatic and asymptomatic PAAs. The PAA DUS images available did not allow us to measure distortion in this study, but we chose to measure diameters and areas of PAAs in different views to better understand anatomic characteristics. In our study, presence or absence of mural thrombus was also significantly more common in PAAs with limb threatening events and symptomatic PAAs. We also found that all largest, AP, and transverse diameters were significantly larger in symptomatic PAAs and PAAs with a limb threatening event compared to asymptomatic PAAs. The smallest patent channel area was in symptomatic PAAs, but the largest thrombus area was in PAAs with a limb threatening event. Creating a percent thrombus burden allowed us to consider the proportion of thrombus burden relative to size, and PAAs with a limb threatening event had a significantly larger burden at 80% than asymptomatic PAAs at 50%. These differences support that size should not be the only criteria considered when defining PAA as high risk.

Given these significant associations with symptom status, we tested the ability of these anatomic measurements to predict either TE or a limb threatening event related to PAAs. In a multivariate model controlling for age, largest diameter was not significantly associated with limb threatening events or TE. Although increasing thrombus size is logically associated with increasing risk of embolism in PAAs, this has never been demonstrated in the literature, possibly due to the absence of a large database with associated ultrasound images. In our database with associated imaging, we have shown that PAAs with a higher percent thrombus burden were 18 times more likely to experience a limb threatening event and 24 times more likely to experience a TE. This is strong evidence in favor of systematically processing DUS data to calculate and list the percent thrombus in every DUS report. We next identified cut point values that had the best discrimination for both TE and limb threatening events for all anatomic characteristics. Percent thrombus at 62% had a better sensitivity (81.3%) and specificity (52%) compared to a diameter of 20.4 mm (sensitivity of 78.1% and specificity of 40.5%) for identifying TEs. For limb threatening events, a diameter of 27.9 mm had a higher specificity than percent thrombus at 73% for differentiating limb threatening events (65.7% vs 62.2%), but percent thrombus had a much higher sensitivity at 69.7% compared to 25.0%. Kaplan Meier curves in Figures 2 and 3 highlight the improved ability of percent thrombus to discriminate between TEs and limb threatening events compared to diameter. These analyses overall support the use of percent thrombus burden in clinical decision making for operative repair of PAAs. One study measuring distortion in PAAs was able to achieve a higher sensitivity of 90% and higher specificity of 89% when using a diameter more than 30 mm with greater than 45° distortion, but this study differentiated between asymptomatic VS symptomatic PAAs and had a smaller sample size of 116 PAAs that did not capture any complications of rupture. In addition, claudication is included in their symptomatic group, which does not necessarily confer increased risk of a limb threatening event.

### Limitations

There are several limitations of the current study. It was difficult to identify a single indication for operative repair of PAAs, as the decision to operate is often multifactorial. The Massachusetts General Brigham multi-institutional retrospective repository is comprised of tertiary hospitals from the Boston area, lending itself to institutional selection and referral biases. Specifically, PAAs with complications from the Boston area may have been referred to and subsequently followed at these tertiary hospitals, limiting the generalizability of this data. The technical details of the DUS performed are unknown and we cannot be sure the same techniques were used to yield consistent results across centers given the retrospective nature of this study. Although compromised distal runoff is noted as criteria for repair of a high risk PAA, we were not able to ascertain runoff based on the DUS images available in this study. Further studies could work on identifying distal runoff to calculate associated predictive values.

## CONCLUSIONS

Clinical practice guidelines for PAAs report a diameter greater than 20 mm should undergo operative repair, but there is little evidence to support this threshold and other anatomic characteristics have not been investigated for predictive value of adverse events related to PAAs. In this study, we created a calculated percent thrombus by accounting for both thrombus area and patent channel area easily measured on DUS. Percent thrombus was significantly associated with both TEs and limb threatening events, whereas diameter was not significantly associated with TEs or limb threatening events. We did confirm that PAA diameter greater than 20.4 mm was predictive of a TE (sensitivity 78.1%, specificity 40.5%) and limb threatening events (sensitivity of 25%, specificity 65.7%). However, percent thrombus greater than 62% outperformed diameter as a predictor of a TE (sensitivity of 81.3%, specificity 52.0%). This analysis supports the use of size greater than 20.4 mm and 62% thrombus in identifying high risk PAAs that warrant repair.

## Data Availability

Data that were used in this study are available on request from the senior author (A.D.). Code to perform analyses in this manuscript are available from the authors upon request (A.D.).

## REFERENCES

1. Pulli R, Dorigo W, Troisi N, Innocenti AA, Pratesi G, Azas L, Pratesi C. Surgical management of popliteal artery aneurysms: Which factors affect outcomes? 2006.

2. Jung G, Leinweber ME, Karl T, Geisbüsch P, Balzer K, Schmandra T, Dietrich T, Derwich W, Gray D, Schmitz-Rixen T, Storck M, Kugelmann U, Schneider C, Engelhardt M, Petzold M, et al. Real-world data of popliteal artery aneurysm treatment: Analysis of the POPART registry. Journal of Vascular Surgery. 2022;75(5):1707–1717.e2.

3. Kim TI, Sumpio BE. Management of Asymptomatic Popliteal Artery Aneurysms. *The International Journal of Angiology : Official Publication of the International College of Angiology*, Inc. 2019;28(1):5.

4. Grip O, Mani K, Altreuther M, Bastos Gonçalves F, Beiles B, Cassar K, Davidovic L, Eldrup N, Lattmann T, Laxdal E, Menyhei G, Setacci C, Settembre N, Thomson I, Venermo M, et al. Contemporary Treatment of Popliteal Artery Aneurysms in 14 Countries: A Vascunet Report. European journal of vascular and endovascular surgery : the official journal of the European Society for Vascular Surgery. 2020;60(5):721–729.

5. Cervin A, Ravn H, Björck M. Ruptured popliteal artery aneurysm. British Journal of Surgery. 2018;105(13):1753–1758.

6. Grip O, Mani K, Altreuther M, Gonçalves FB, Beiles B, Cassar K, Davidovic L, Eldrup N, Lattmann T, Laxdal E, Menyhei G, Setacci C, Settembre N, Thomson I, Venermo M, et al. Contemporary Treatment of Popliteal Artery Aneurysms in 14 Countries: A Vascunet Report.

7. Pulli R, Dorigo W, Troisi N, Innocenti AA, Pratesi G, Azas L, Pratesi C. Surgical management of popliteal artery aneurysms: Which factors affect outcomes? 2006.

8. Kim TI, Sumpio BE. Management of Asymptomatic Popliteal Artery Aneurysms. The International Journal of Angiology : Official Publication of the International College of Angiology, Inc. 2019;28(1):5.

9. Farber A. Surgery appears to outperform endovascular therapy for popliteal artery aneurysms; however, the real answer as to which treatment strategy works best and for whom remains elusive. Journal of vascular surgery. 2022;75(5):1718–1719.

10. Lowell RC, Gloviczki P, Hallett JW, Naessens JM, Maus TP, Cherry KJ, Bower TC, Pairolero PC. Popliteal artery aneurysms: the risk of nonoperative management. Annals of vascular surgery. 1994;8(1):14–23.

11. Mahmood A, Salaman R, Sintler M, Smith SRG, Simms MH, Vohra RK. Surgery of popliteal artery aneurysms: A 12-year experience. Journal of Vascular Surgery. 2003;37(3):586– 593.

12. Jergovic I, Cheesman MA, Siika A, Khashram M, Paris SM, Roy J, Hultgren R. Natural history, growth rates, and treatment of popliteal artery aneurysms. Journal of vascular surgery. 2022;75(1):205–212.e3.

13. Del Tatto B, Lejay A, Meteyer V, Roussin M, Georg Y, Thaveau F, Geny B, Chakfe N. Open and Endovascular Repair of Popliteal Artery Aneurysms. Annals of Vascular Surgery. 2018;50:119–127.

14. Ascher E, Markevich N, Schutzer RW, Kallakuri S, Jacob T, Hingorani AP. Small popliteal artery aneurysms: Are they clinically significant? Journal of Vascular Surgery. 2003;37(4):755–760.

15. Kropman RHJ, van Santvoort HC, Teijink J, van de Pavoordt HDWM, Belgers HJ, Moll FL, de Vries JPPM. The medial versus the posterior approach in the repair of popliteal artery aneurysms: A multicenter case-matched study. Journal of Vascular Surgery. 2007;46(1):24–30.

16. Galland RB, Magee TR. Popliteal aneurysms: distortion and size related to symptoms. European journal of vascular and endovascular surgery : the official journal of the European Society for Vascular Surgery. 2005;30(5):534–538.

17. Farber A, Angle N, Avgerinos E, Dubois L, Eslami M, Geraghty P, Haurani M, Jim J, Ketteler E, Pulli R, Siracuse JJ, Murad MH. The Society for Vascular Surgery clinical practice guidelines on popliteal artery aneurysms. Journal of Vascular Surgery. 2022;75(1):109S–120S.

18. Isselbacher EM, Preventza O, Black JH, Augoustides JG, Beck AW, Bolen MA, Braverman AC, Bray BE, Brown-Zimmerman MM, Chen EP, Collins TJ, DeAnda A, Fanola CL, Girardi LN, Hicks CW, et al. 2022 ACC/AHA Guideline for the Diagnosis and Management of Aortic Disease: A Report of the American Heart Association/American College of Cardiology Joint Committee on Clinical Practice Guidelines. Circulation. 2022;146(24):E334–E482.

19. Chaikof EL, Dalman RL, Eskandari MK, Jackson BM, Lee WA, Mansour MA, Mastracci TM, Mell M, Murad MH, Nguyen LL, Oderich GS, Patel MS, Schermerhorn ML, Starnes BW. The Society for Vascular Surgery practice guidelines on the care of patients with an abdominal aortic aneurysm. Journal of Vascular Surgery. 2018;67(1):2–77.e2.

20. Chaer RA, Abularrage CJ, Coleman DM, Eslami MH, Kashyap VS, Rockman C, Murad MH. The Society for Vascular Surgery clinical practice guidelines on the management of visceral aneurysms. Journal of Vascular Surgery. 2020;72(1):3S–39S.

21. Steenberge SP, Caputo FJ, Rowse JW, Lyden SP, Quatromoni JG, Kirksey L, Smolock CJ. Natural history and growth rates of isolated common iliac artery aneurysms. Journal of Vascular Surgery. 2022;76(2):461–465.

22. Sweeting MJ, Thompson SG, Brown LC, Powell JT. Meta-analysis of individual patient data to examine factors affecting growth and rupture of small abdominal aortic aneurysms. The British journal of surgery. 2012;99(5):655–665.

23. Davies RR, Goldstein LJ, Coady MA, Tittle SL, Rizzo JA, Kopf GS, Elefteriades JA. Yearly rupture or dissection rates for thoracic aortic aneurysms: simple prediction based on size. The Annals of Thoracic Surgery. 2002;73(1):17–28.

24. Tomaszewski KA, Popieluszko P, Graves MJ, Pękala PA, Henry BM, Roy J, Hsieh WC, Walocha JA. The evidence-based surgical anatomy of the popliteal artery and the variations in its branching patterns. Journal of Vascular Surgery. 2017;65(2):521–529.e6.

25. Dorigo W, Pulli R, Turini F, Pratesi G, Credi G, Innocenti AA, Pratesi C. Acute Leg Ischaemia from Thrombosed Popliteal Artery Aneurysms: Role of Preoperative Thrombolysis. European Journal of Vascular and Endovascular Surgery. 2002;23(3):251–254.

26. Jung G, Leinweber ME, Karl T, Geisbüsch P, Balzer K, Schmandra T, Dietrich T, Derwich W, Gray D, Schmitz-Rixen T, Storck M, Kugelmann U, Schneider C, Engelhardt M, Petzold M, et al. Real-world data of popliteal artery aneurysm treatment: Analysis of the POPART registry. Journal of vascular surgery. 2022;75(5):1707–1717.e2.

27. Ascher E, Markevich N, Schutzer RW, Kallakuri S, Jacob T, Hingorani AP. Small popliteal artery aneurysms: Are they clinically significant? Journal of Vascular Surgery. 2003;37(4):755–760.

28. Ribeiro M, Oderich GS, Macedo T, Vrtiska TJ, Hofer J, Chini J, Mendes B, Cha S. Assessment of aortic wall thrombus predicts outcomes of endovascular repair of complex aortic aneurysms using fenestrated and branched endografts. Journal of vascular surgery. 2017;66(5):1321–1333.

